# Accuracy of the Oxford Visual Perception Screen

**DOI:** 10.1101/2025.06.04.25328952

**Authors:** Kate Cowen, Faye Tabone, Sam Webb, Andrea Kusec, Ruth DaSilva, Revin Thomas, Nele Demeyere, Lisa Shaw, Kathleen Vancleef

**Affiliations:** Department of Psychology, Durham University; Nuffield Department of Clinical Neurosciences, University of Oxford; Gateshead Health NHS Foundation Trust; Country Durham and Darlington NHS Foundation Trust; Population Health Sciences Institute, Newcastle University

**Keywords:** stroke, visual perception, diagnostic accuracy, sensitivity, specificity, screening

## Abstract

**Background:** Post-stroke visual perception difficulties are common, though not always screened for given limited time and available assessments. The aim is to evaluate diagnostic accuracy of a new short assessment, the Oxford Visual Perception Screen (OxVPS) which has been developed to support clinicians in standardised assessment of visual perception difficulties in line with clinical guidelines.

**Methods:** In a cross-sectional prospective study, stroke survivors were recruited across three subacute rehabilitation units in the United Kingdom between June 2023 and March 2024. Inclusion criteria were 18 years and older, diagnosis of stroke within six weeks, able to concentrate for 15 minutes, and understand simple English instructions. Participants completed the index test, OxVPS, and reference standard, the Rivermead Perceptual Assessment Battery (RPAB) within two weeks. Diagnostic accuracy was determined via a Receiver Operator Curve analysis of complete cases which included estimation of the Area Under the Curve (AUC) and of sensitivity and specificity at the optimal cut-off point (Youden Index J). A secondary analysis was performed on all cases in which missing data were replaced by random forest imputations. Completion time and rate were compared with a t-test and Wilcoxon test.

**Results:** The AUC for complete cases (n=33) was estimated at 0.83 with a 95% confidence interval ranging from 0.68 to 0.98. At the optimal OxVPS cut-off point of 2, sensitivity was estimated at 85% (95% CI: 0.54-1) and specificity at 77% (95% CI: 0.54-1). The analysis on all data (n=62) estimated AUC at 0.88 (95% CI=0.80-0.96), sensitivity at 87% (95% CI=0.62-1) and specificity at 80% (95% CI=0.67-1). A shorter completion time (13 min, t(32)=-20.08, p<0.001) and higher completion rate (97%, V=118, p=0.03) of OxVPS compared to RPAB.

**Conclusions:** Our results indicate that OxVPS is a time-efficient assessment suitable for time-limited clinical settings that can correctly identify stroke survivors with and without visual perception difficulties.

## Background

Visual problems are common post stroke with prevalence estimates ranging from 20.5% to 76%, inclusive of visual perception difficulties[1, 2, 3]. Visual perception is the dynamic process of perceiving the environment through reception of sensory inputs and translating them into meaningful concepts associated with visual knowledge of the environment [4]. Examples of difficulties in visual perception include struggling to recognise objects (associative agnosia), and/or faces (prosopagnosia), or difficulty reading (alexia).

Visual perception difficulties can impact stroke survivors’ quality of life [5], functional outcomes [6], participation, independence and pose substantial risks (e.g., participation in traffic, cooking risks) [5,6]. Rehabilitation for visual perception difficulties can include training (e.g., online reading training), adaptations to the environment (e.g., remove distractions), or coping strategies (e.g., use memory or other senses) [7]. Improved identification (e.g., screening) can support care planning [8], reduce the need for additional care (e.g., supporting the performance of activities of daily living), avoid further medical issues (e.g., falls), and improve mental health (e.g., more social interactions) [9].

Standardised assessment instruments for visual perception difficulties in stroke survivors, such as, the Rivermead Perceptual Assessment Battery (RPAB) and the Visual Object and Space Perception Battery are too time consuming (40-120 minutes) for screening in clinical practice, unsuitable for patients with communication difficulties and assess only a limited range of visual perceptual functions [10]. A recent survey reported that 94% of clinicians rely on patient self-report and clinical observation for the diagnosis of visual perception difficulties [11]. However, a systematic review demonstrated that the sensitivity of screening is substantially lowered in patients who are unable to report their symptoms [8]. National clinical guidelines also recommend the use of standardised assessments in evaluations visual perception difficulties after stroke [12]. Additionally, research with 214 clinicians has confirmed the need for a quick, evidence-based screening tool suitable for stroke survivors which can identify a range of visual perception difficulties [11,13].

In response to this clinical need, the Oxford Visual Perception Screen (OxVPS) has been developed [14]. The OxVPS is a paper-and-pencil test which screens for 15 visual perception difficulties in under 15 minutes through 10 subtests. The subtests include tasks such as picture naming, item counting, and a figure copy which has been standardised using normative data in neurologically healthy volunteers [14]. The OxVPS aspires to support clinicians’ practice by resolving some of the challenges clinicians face with current standardised assessment instruments. Demonstrating diagnostic accuracy to detect visual perception impairments is essential before OxVPS can be used by clinicians to assess stroke patients for visual perception difficulties in sub-acute and rehabilitation settings.

## Aim

The aim of the study was to evaluate the diagnostic accuracy of the OxVPS in detecting visual perception difficulties in stroke survivors.

## Methods

### Participants

This was a prospective cross-sectional study which conforms to STARD guidelines in its design and reporting [15]. A convenience sample of stroke survivors were recruited at three stroke rehabilitation units in the North East and in the South East of England, United Kingdom, between June 2023 and March 2024. The inclusion criteria were: 18 years or older and a clinical diagnosis of stroke (ischemic stroke and/or intracerebral haemorrhage) in the past six weeks. Patients were excluded if they had insufficient understanding of English, no capacity to provide informed consent, or were unable to follow instructions or concentrate for 15 minutes. All patients admitted with a stroke at each participating site were systematically screened against the study eligibility criteria by trained members of the clinical multidisciplinary team. Consent for study procedures and data collection were subsequently sought by a member of the research team.

### Instruments

#### Index test

The index test, the Oxford Visual Perception Screen (OxVPS) [14], screens for visual perception difficulties following stroke. OxVPS meets clinical requirements by being quick to learn, easy to administer (minimal training), and suitable for stroke survivors with communication difficulties [14]. The OxVPS is described in detail in Vancleef et al, 2025 [14].

### Reference standard

The Rivermead Perceptual Assessment Battery (RPAB) [16] is a standardised assessment developed for assessing and quantifying deficits in visual perception following a stroke. In 45-120 minutes, RPAB evaluates eight visual functional categories across 16 subtests: form constancy, colour constancy, sequencing, object completion, figure ground discrimination, body image, spatial awareness, and inattention.

The clinical usefulness of the RPAB was established since its inception in 1985 [16] and is currently still the most widely used assessment tool for visual perception difficulties in the United Kingdom and Ireland [11]. All subtests demonstrate excellent inter-rater reliability (correlations between 0.72 and 1, >0.90 in 14 out of 16 subtests) and fair test-retest reliability (correlations between 0.27 and 1, >0.70 in 10 out of 16 subtests) [16]. Concurrent validity was established by comparing scores of patients with a brain injury and healthy volunteers and except for one subtest (Series: measuring form constancy and object matching), the scores between both groups are significantly different. The effect of group (stroke) explained between 3% to 23% of the variance in scores on each of the subtests [17]. Furthermore, Friedman and Leong [18] found that RPAB is a significant predictor of functional outcomes at six months as measured with the Barthel index (r=0.4) and a better predictor than arm strength, hemisensory loss, hemianopia, age, or visual inattention suggesting strong predictive validity. The clinical utility and the good psychometric properties of RPAB made it an appropriate choice as a reference standard.

### Procedures

The research team were guided by a steering committee (academics, medical doctors, occupational therapists and stroke survivors) and Patient and Public Involvement panel to establish appropriate and acceptable procedures. There was a 2-week maximum limit observed between OxVPS and RPAB to avoid performance changes due to potential recovery [8, 20] and the tests were administered in no particular order based on logistics. For example, room availability (RPAB difficult to administer bedside), participant energy level, time constraints of researcher and participant (patient’s therapy/visitors) and other practical considerations when collecting data on a busy ward. To reduce any bias, research team members administering OxVPS were blinded to the outcome of RPAB and vice versa. In addition, the research team only collected demographic information (age, gender, ethnicity, etc.) and details about a participant’s stroke (e.g., type, severity, CT scan, location) after completion of the assessments.

### Sample Size

The sample size was determined based on the pre-specified Receiving Operator Curve (ROC) analysis with a probability of a Type 1 error of 0.05 and minimum power of 0.90. The minimum Area Under the Curve (AUC) that would make OxVPS clinically meaningful was set at 0.80 which is generally considered excellent [21]. Previous research into the frequency of visual perception difficulties in the first month after stroke range from 20.5% to 76% [2, 3]. Sensitivity analyses showed that the required sample size varied between 34 (for prevalence of 50%) and 54 (for prevalence of 20.5%, a prevalence of 76% required a sample size of 45). Based on these calculations, the minimum sample size was set at 54. A high attrition was expected because of the long duration (120 minutes) of RPAB, the need to complete both assessments within two weeks, and conflicts between participants’ therapy schedule and availability of researchers on the stroke unit. Therefore, over-recruitment was intentional. Note that the study was not powered for sensitivity or specificity.

### Analysis

In a pre-specified analysis including only complete cases, the AUC of the ROC was constructed empirically, and 95% confidence intervals were calculated with the DeLong method [22]. The optimal cut-off point for OxVPS was determined through Youden Index J for which the sensitivity, specificity, positive and negative predictive values was calculated. The 95% confidence intervals of these four values were calculated through bootstrapping with 10,000 iterations.

Inspection of the data revealed that a small proportion of missing data in RPAB or OxVPS per participant was common. Hence, an exploratory secondary analysis was conducted for which the missing subtest scores of RPAB and OxVPS were replaced by random forest imputation. In this secondary analysis, a modified Youden index was calculated in which sensitivity was given up to four times more weight than specificity (compared to equal weight in the traditional Youden index) because for a screening test high sensitivity (few false negatives) is more important than high specificity (few false positives) [23]. A sensitivity analysis further explored the effect of different weights for sensitivity and specificity.

## Results

Throughout recruitment, as seen in Figure 1, 766 stroke patients were screened against the inclusion criteria, of which 250 were eligible for participation in the study. Out of these, 116 engaged with test administration of the OxVPS (index test). However, 54 out of the 116 participants were not able to engage with the RPAB for various reasons such as fatigue, discharge, participating in therapy, or having visitors when the researchers were on the stroke unit. This meant 62 out of 116 took part in both OxVPS (index test) and RPAB (reference standard). Of these 62 participants, 26 (42%) completed OxVPS before RPAB, while 36 (58%) completed RPAB before OxVPS. The average time interval between OxVPS and RPAB was 6.34 days (SD=3.96).

**Figure 1:**
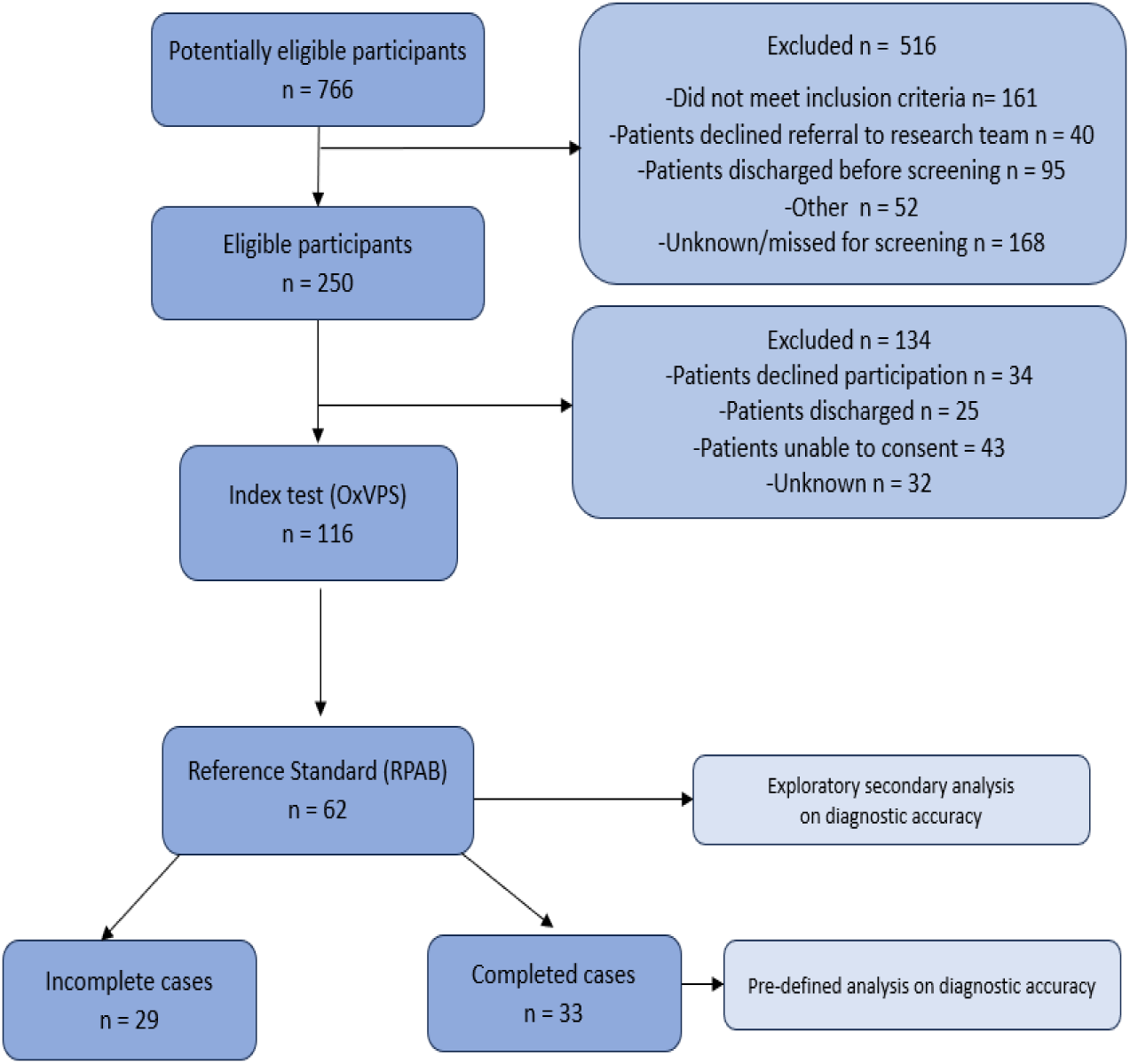
Participant flow chart of all participants who took part in the study. Not all participants completed all subtests of OxVPS and RPAB. Two analyses were performed: a pre-defined analysis on the complete cases only (n=33) and an exploratory secondary analysis on all cases (n=62).

Forty-seven percent of participants (n=29) could not complete all subtests of OxVPS and RPAB due to aphasia, fatigue, inability to hold a pen, or difficulties understanding the instructions. This was more common for the RPAB than for the OxVPS: 26 did not complete RPAB and 10 did not complete OxVPS. The average percentage of incomplete RPAB subtests was 4.7% and significantly higher than the average percentage of incomplete OxVPS subtests which was 2.98% (Wilcoxon test, V=118, p=0.03). Further details about missing data are provided in the Supplementary materials.

Reports of baseline demographic and clinical characteristics of participants for the whole sample and both subsamples of complete (n=33) and incomplete cases (n= 62) can be viewed in Table 1. To avoid inflating the Type I error, only statistical tests for the difference between subsamples on the key outcome: visual perception difficulties were performed.

**Table 1:** Participant baseline demographics.

| Variables | All participants<br>(n=62) | Complete<br>cases (n=33) | Incomplete<br>cases (n=29) |
| --- | --- | --- | --- |
| Age in years [*mean (SD)*] | 74.13 (12.43) | 74.19 (13.68) | 74.05 (11.08) |
| Gender [*count (percentage)*] |  |  |  |
| Female | 34 (55%) | 20 (61%) | 14 (48%) |
| Male | 28 (45%) | 13 (39%) | 15 (52%) |
| Ethnicity [*count (percentage)*] |  |  |  |
| White | 61 (98%) | 32 (97%) | 29 (100%) |
| Other | 1 (2%) | 1 (3%) | 0 (0%) |
| Handedness |  |  |  |
| Right-handed | 59 (95%) | 33 (100%) | 26 (90%) |
| Left-handed | 3 (5%) | 0 (0%) | 3 (10%) |
| Qualifications [*count (percentage)*] |  |  |  |
| Information not available | 24 (39%) | 11 (33%) | 13 (45%) |
| No qualifications | 8 (13%) | 5 (15%) | 3 (10%) |
| GCSE/CSE | 16 (26%) | 12 (36%) | 4 (14%) |
| A /O Level | 4 (6%) | 2 (6%) | 2 (7%) |
| University | 2 (3%) | 0 (0%) | 2 (7%) |
| Other | 8 (13%) | 3 (9%) | 5 (17%) |
| Days since stroke [*mean (SD)*] | 28.74 (10.94) | 27.97 (10.59) | 29.62 (11.44) |
| Stroke severity [*count (percentage)*] |  |  |  |
| Minor (NIHSS 1-4) | 20 (32%) | 14 (42%) | 6 (21%) |
| Moderate (NIHSS 5-15) | 35 (56%) | 16 (48%) | 19 (66%) |
| Severe (NIHSS 16-20) | 4 (6%) | 3 (9%) | 1 (3%) |
| Information not available | 3 (5%) | 0 (0%) | 3 (10%) |
| Type of stroke [*count (percentage)*] |  |  |  |
| Ischemic | 46 (74%) | 27 (82%) | 19 (66%) |
| Haemorrhagic | 16 (26%) | 6 (18%) | 10 (34%) |
| Oxford stroke classification [*count (percentage)*] |  |  |  |
| TACS | 15 (24%) | 5 (15%) | 10 (34%) |
| PACS | 22 (35%) | 14 (42%) | 8 (28%) |
| POCS | 6 (10%) | 4 (12%) | 2 (7%) |
| LACS | 19 (31%) | 10 (30%) | 9 (31%) |
| Side of stroke *[count (percentage)]* |  |  |  |
| Left | 19 (31%) | 11 (33%) | 8 (28%) |
| Right | 41 (66%) | 20 (61%) | 21 (72%) |
| Bilateral | 2 (3%) | 2 (6%) | 0 (0%) |
| Other neurological conditions<br>*[count (percentage)]* |  |  |  |
| None | 50 (81%) | 28 (85%) | 22 (76%) |
| Other | 12 (19%) | 5 (15%) | 7 (24%) |

GCSE: General Certificate of Secondary Education; CSE: Certificate of Secondary Education; A-level: Advanced Level qualification; O-Level: Ordinary Level qualification; NIHSS: National Institute of Health Stroke Scale; TACS: Total Anterior Circulation Stroke; PACS: Partial Anterior Circulation Stroke; POCS: Posterior Circulation Syndrome; LACS: Lacunar Stroke.

Participants who completed all subtests had on average higher RPAB scores (mean number of subtests passed=11) than participants who did not complete all subtests (mean number of subtests passed=6.9, 95% confidence interval for difference=[2.27, 5.94], t(51.74)=4.49, p<0.001, Figure 2). The same pattern was observed for scores on OxVPS (95% confidence interval for difference=[1.28, 3.73], t(52.81)=4.10, p < 0.001). This suggests that the sample of participants who completed all subtests is biased towards participants with less severe visual perception difficulties. Across the whole sample, severity of visual perception difficulties ranged from very severe (0 out of 16 subtests passed on the RPAB) to negligible (a score of 15 out of 16 on RPAB) with an average score on RPAB of 9.08 (SD=4.06).

**Figure 2.**
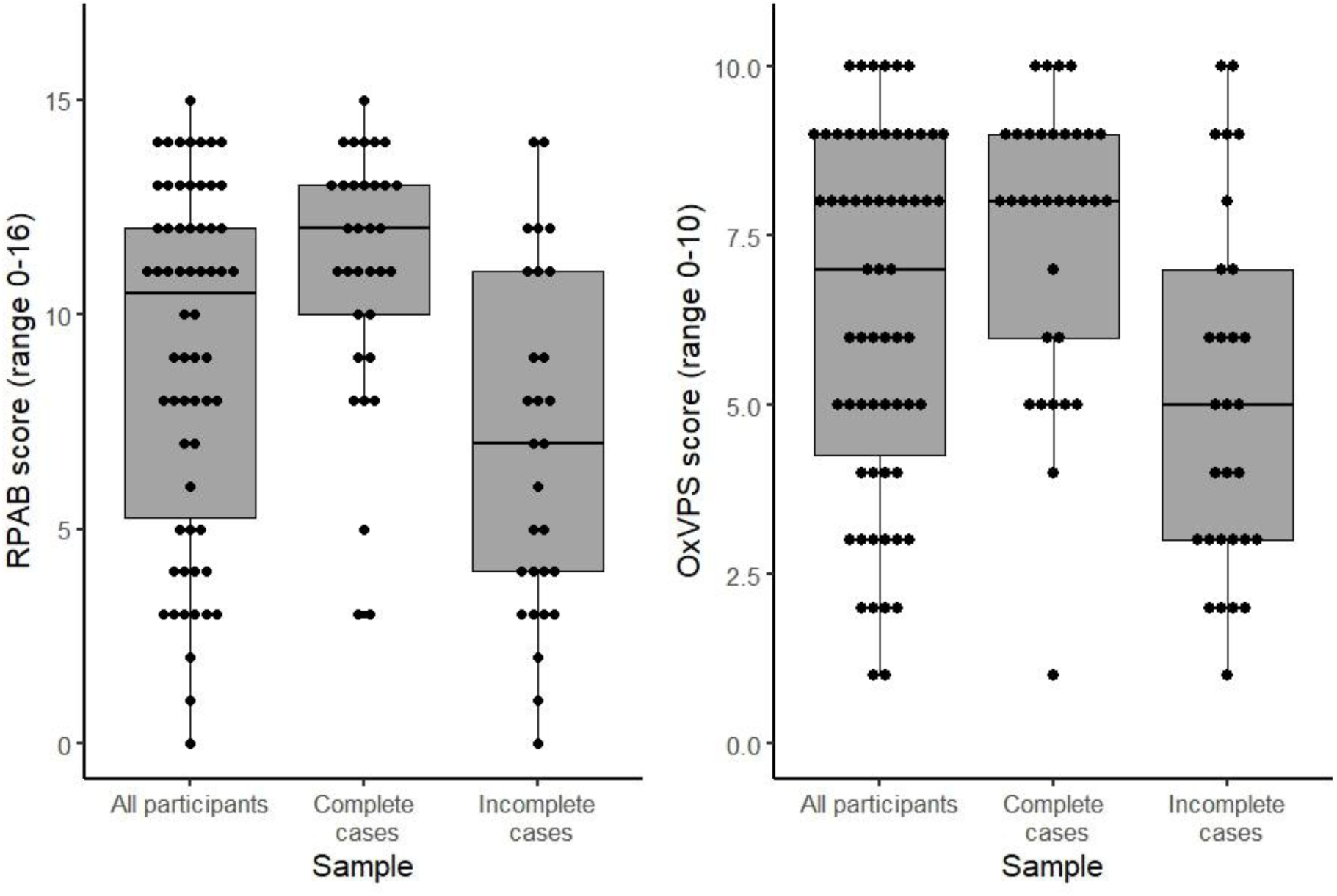
Scores on RPAB (left panel) and OxVPS (right panel) for the full sample (n=62) and subsamples (n=33 and n=29). Performance is expressed as a criterion score of passed subtests. A passed subtest is a subtest with a score above the cut-off value for normal performance based on a sample of healthy volunteers. Therefore, a higher score means better visual perception. Boxplots for each (sub)sample show the median score as the horizontal line inside the box and first and third quantile as the lower and upper hinge of the box. The whiskers extend from the hinge to the highest/lowest value no further than 1.5 times the interquartile range from the hinge. All individual data points are superimposed on the boxplots. In the left boxplot of each panel, missing data on a subtest were replaced by random forest imputations.

### Diagnostic accuracy

The estimated AUC in the ROC analysis on complete cases (n=33, pre-specified analysis) was 0.83 (Figure 3) with the 95% confidence interval ranging from 0.68 to 0.98. The Youden Index indicated an optimal cut-off point for visual perception difficulties at two or more failed subtests on OxVPS. At this cut-off, OxVPS has an estimated sensitivity of 0.85 (95% confidence interval 0.54 - 1), an estimated specificity of 0.77 (95% confidence interval 0.54 -1), an estimated positive predictive value of 0.85 (95% confidence interval 0.68 - 1) and an estimated negative predictive value of 0.77 (95% confidence interval 0.48 - 1, Table 2,). Note that the study was powered for AUC and not for sensitivity, specificity, positive or negative predictive value hence the wide confidence intervals for these measures.

**Figure 3.**
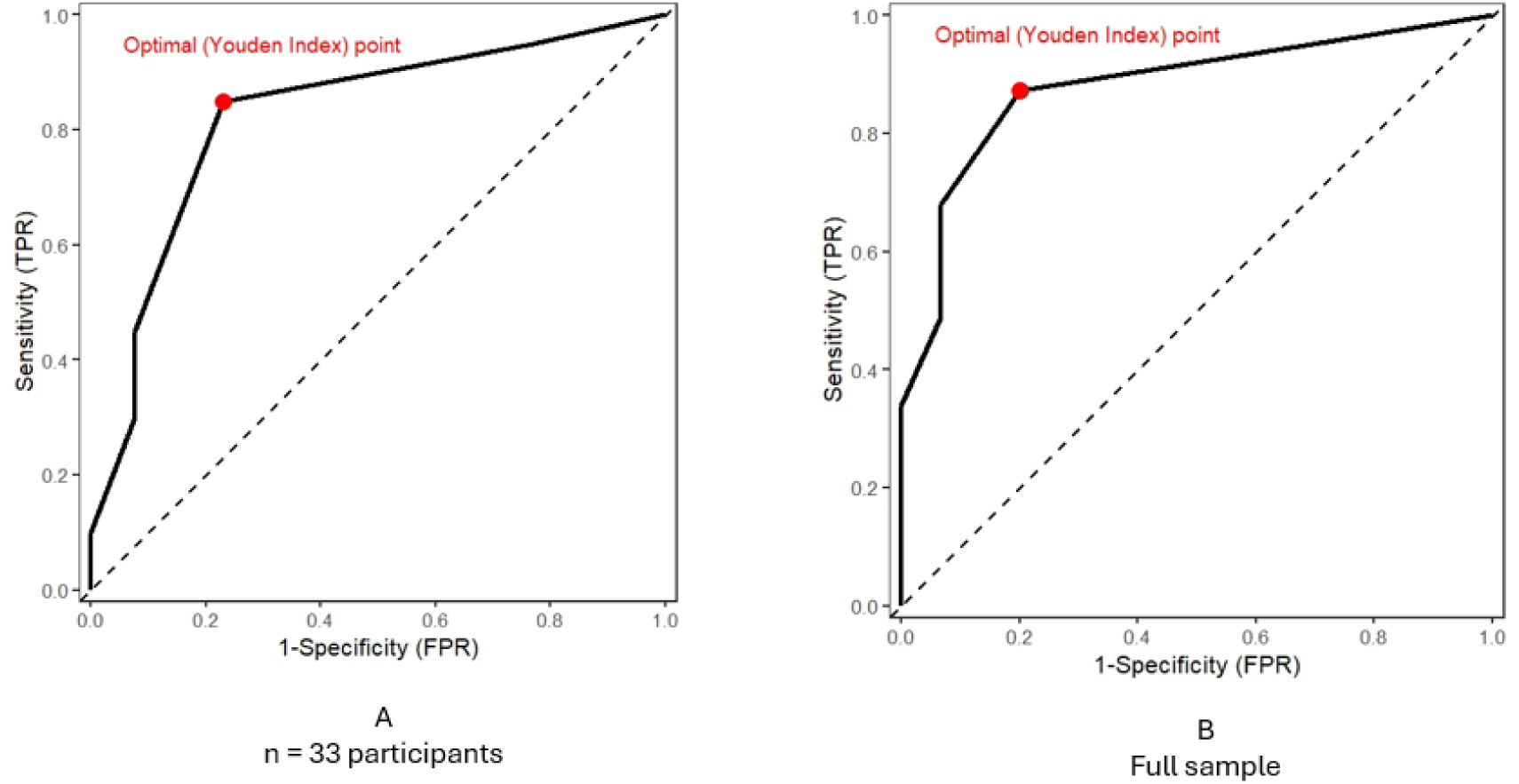
Receiver Operating Curve of OxVPS for the detection of visual perception difficulties as indicated by the RPAB in the sample of participants who completed all subtests panel A, and the full sample, panel B. The solid line represents the empirical ROC. The dashed line shows the chance line. At the optimal Youden index point (dot). In panel A the sensitivity is estimate at 0.85 and the specificity at 0.77, in panel B the sensitivity is estimate at 0.87 and the specificity at 0.80.

**Table 2.** Accuracy of OxVPS for screening of visual perception difficulties in the sample of participants who completed all subtests.

| OxVPS results | RPAB results |  | Total |
| --- | --- | --- | --- |
|  | Visual perception difficulties | No visual perception difficulties |  |
| Visual perception difficulties | 17 | 3 | 20 |
| No visual perception difficulties | 3 | 10 | 13 |
| Total | 20 | 13 | 33 |

The exploratory secondary analyses on all data (n=62) estimated the AUC at 0.88 (Figure 3) with a 95% confidence interval ranging from 0.80 to 0.96. The modified Youden index which puts more weight on sensitivity compared to specificity, indicated an optimal cut-off point at two or more failed subtests on OxVPS. At this cut-off point, sensitivity is estimated at 0.87 (95% confidence interval: 0.62 - 1), specificity at 0.80 (95% confidence interval: 0.67 - 1), positive predictive value at 0.93 (95% confidence interval: 0.88 - 1), and negative predictive value at 0.67 (95% confidence interval: 0.37 - 0.88, Table 3). Sensitivity analyses revealed that varying the weights for sensitivity and specificity did not change these results as long as sensitivity was given at least equal weight as specificity (see Supplementary materials).

**Table 3.** Accuracy of OxVPS for screening of visual perception difficulties for the full sample.

| OxVPS results | RPAB results |  | Total |
| --- | --- | --- | --- |
|  | Visual perception difficulties | No visual perception difficulties |  |
| Visual perception difficulties | 41 | 3 | 44 |
| No visual perception difficulties | 6 | 12 | 18 |
| Total | 47 | 15 | 62 |

### Completion time and adverse events

The average time to complete OxVPS (13 minutes) was significantly shorter than the average time to complete the RPAB (53 minutes, t(32)=-20.08, p < 0.001, complete cases only). In conjunction with the data on missingness, this suggests that compared to the RPAB, patients are more likely to complete OxVPS and within a shorter time. There were no adverse events from participants completing either the reference standard or index test.

## Discussion

This study evaluated the diagnostic accuracy of the OxVPS, a novel screening tool for visual perception difficulties following stroke. Participants’ performance on the OxVPS and RPAB were closely related. The results of the pre-defined analysis in a subsample completing all OxVPS and RPAB subtests suggest good sensitivity [23], with the OxVPS identifying 85% of participants with visual perception difficulties and good specificity, categorising 77% of participants without visual perception difficulties correctly [24]. However, the wide confidence intervals recommend cautious interpretation. In an exploratory secondary analysis of the whole sample sensitivity increased to 87% and specificity to 80%. Although defined post-hoc, this analysis has higher external validity because it includes participants with more severe strokes and more severe visual perception difficulties. In addition, the confidence intervals are narrower because of the larger sample size.

The results of the current study are comparable with other visual perception screening tools which have undergone similar accuracy studies with stroke survivors. For example, the five subdomain scores of the Occupational Therapy Assessment for Perceptual Screening Test had estimated sensitivity ranging from 30.8% to 90.4% and specificity ranging from 68% to 95.7% against the Lowenstein Occupational Therapy Assessment standard and geriatric version as the reference standard [25].

In the full sample, the prevalence of participants found to have visual perception problems as identified by the RPAB was 76%. This is in line with earlier reports using RPAB in a similar population [2], but in stark contrast to more recent reports with Rowe et al. [3] finding only 20.5% of stroke survivors had visual perception issues following stroke. Rowe et al. [3] relied on self-report and observations in functional tasks except when assessing inattention (using cancellation/line bisection tests) which better reflects current clinical practice in the UK [11]. In contrast, both the RPAB and the OxVPS asses a range of visual perception difficulties through standardised performance-based assessments with cut-off scores supported by normative data. This suggests that standard assessments and indeed the OxVPS are able to pick-up more subtle visual perception problems that may typically not be reported by patients and therefore missed in standard clinical practice.

The argument of interplay and overlap between visual perception and cognition has been debated for some time [28]. We note that some cognitive penetration within visual perception always occurs [29], and the impact of cognition becomes more profound at later stages of the visual perception process [30]. Some of the tasks within the OxVPS and RPAB focus on these later stages such as spatial inattention and visuo-motor abilities or praxis. In addition, completion of OxVPS and RPAB relies on sustained attention, organisation and planning [31]. Like many diagnostic accuracy studies in neuropsychology, finding a single gold standard measure that measures a brain function in isolation is often unattainable [26]. Therefore, in accordance with STARD guidelines [27], the best available assessment was used as the reference standard. The longevity, clinician perceptions about usefulness in stroke settings [17,18,20] and the overlap in measurement domains with OxVPS made RPAB a suitable choice.

The impact of cognitive abilities on test performance is also reflected in the recommendation to use different RPAB cut-off scores for different intelligence levels [16]. However, it was not feasible to administer intelligence tests in addition to the OxVPS and RPAB given participants were recovering from a stroke. Therefore, cut-off scores for average intelligence were used. The comparable prevalence of visual perception difficulties in the current sample and in an RPAB study [16] with intelligence-adjusted scores suggests that at group level the choice for average intelligence cut-offs has been acceptable.

The key limitations of the study are the imprecise estimates of sensitivity and specificity, and the lack of education and intelligence measures which may have impacted scores on RPAB and OxVPS [19, 34]. As the first study into diagnostic accuracy this study was aimed at determining a cut-off point for the OxVPS overall score, the study was powered for AUC and not for estimates of sensitivity and specificity. The estimation of sensitivity and specificity are therefore imprecise, especially in the pre-defined analysis which uses a smaller sample. The estimates from the study can be used to plan a follow-up study to determine sensitivity and specificity of OxVPS with higher precision.

The study’s strengths include representativeness of the sample, inclusive recruitment and strategies to minimise bias and increase generalisability. The sample is similar in age, gender, and stroke severity to the Sentinel Stroke National Audit Programme (SSNAP) [35], thus strengthening the evidence that the OxVPS is a feasible assessment in the general stroke population. To minimise bias, the research team were blinded to results of the OxVPS and the RPAB when administering the tests and information from the participant’s medical notes were only recorded after test administrations. Additionally, inclusion of variation of perspectives and expertise (patient and public engagement and the steering committee) has ensured the rigorous and inclusive nature of the study design, prioritised participant experience and empowered stroke survivors [36]. For example, patient and public involvement resulted in accessible forms for patients with communication difficulties which enhanced inclusivity for recruitment, and of study closure at the earliest opportunity given the onerous nature of some RPAB tasks and high prevalence of fatigue following stroke [37].

## Conclusions

The results of the study indicate that the OxVPS can distinguish between stroke survivors with and without visual perception difficulties, suggesting strong clinical utility. Given that most participants were able to complete all subtests of the OxVPS and within 15 minutes, the OxVPS may be a more feasible assessment within the stroke population compared to the more time-intensive reference standard, RPAB and offer a resource to efficiently screen for visual perception difficulties following stroke.

## Data Availability

The dataset and analysis code are available: https://osf.io/2kd7s/?view_only=fb3f27c55a6e4dcbb98cfb8ce923903b

## Acknowledgements

The research team would like to acknowledge and thank the study’s steering committee for their advice and expertise: Hazel Hammond, Steve Darcy, Dr Amanda Ellison, Dr Alison Lane, and Kirsty Forrester. Our gratitude also extends to the NHS principal investigators Emma Garrett, Vivienne Southcott and research nurses, Vivienne Rudge and Beverley McClelland. Furthermore, we appreciate the valuable insight from the North East and Cumbria Stroke patient and carers panel, and the support from research assistants of the Visual Neuropsychology Research Group at Durham University.

## Authors’ contributions

Conceptualisation (KV, ND, LS). Methodology (KC, ND, LS, KV). Validation (KC, ND, LS, KV). Investigation (KC, FT, AK, SW, KV). Resources (KC, RD, RT, KV). Visualisation (KC, KV)

Supervision (KC, KV, RD, RT, ND, KV). Project administration (KC, KV). Funding acquisition (KV, ND, LS). Writing-original draft (KV, KC). Formal analysis & Data Curation (KV)

All co-authors have approved the final version of the manuscript for submission.

## Ethical approval and informed consent statement

Ethical approval was obtained from the Health Research Authority, REC reference 23/EM/0086 by the Derby Research Ethics committee. All participants have provided written informed consent for all aspects of the study, including dissemination.

## Study Registration and Protocol

The trial has been registered with ClinicalTrials.gov: ID NCT05981482 on 27/07/2023. The process was initiated prior to recruitment. However, registration completed following the recruitment of 16 participants

The full study protocol can be accessed at https://clinicaltrials.gov/study/NCT05981482.

## Supplementary materials to The Oxford Visual Perception Screen detects visual perception difficulties in sub-acute adult stroke survivors: a cross-sectional diagnostic accuracy study

### Missing data

In RPAB a value was considered missing and imputed if the participant did not attempt a single trial in the subtest. If they had attempted at least one trial, it was assumed they did not complete the task because the task was too difficult and therefore, they received a score of 0 for the skipped trials. In OxVPS, such trials were clearly labelled as “no answer” and were also assigned a score of 0. Blank responses on OxVPS were considered missing data.

We explored missing data on the subtest scores of OxVPS and RPAB. Not fully completing the OxVPS or the RPAB was common in our sample. Only 53% completed both assessments.

Missingness was more common for RPAB than for OxVPS, with 42% (26 participants) not completing RPAB versus only 16% (10 participants) not completing OxVPS. However, missingness per participant was low: on average only 3.23% of OxVPS subtest scores were missing for a given participant and 4.75% of RPAB subtest scores. The breakdown in Table S1 shows that most participants have 0% missing subtest scores. Of the people with missing data, most missed less than 20% of subtest scores, with only one participant missing more than 50% of the OxVPS subtest scores.

**Table S1.** Number of participants per percentage missing subtest scores on RPAB or OxVPS.

| % missing | 0 | 1-10 | 11-20 | 21-30 | 31-40 | 41-50 | 51-60 | 61-70 |
| --- | --- | --- | --- | --- | --- | --- | --- | --- |
| RPAB | 36 | 16 | 6 | 2 | 2 | 0 | 0 | 0 |
| OxVPS | 52 | 5 | 2 | 1 | 1 | 0 | 0 | 1 |

Viewed per subtest, on average, 1.45% of scores on a subtest were missing with a maximum of 21.54% of missing scores for the Left Right Copying Words subtest of the RPAB. Further details are given in Table S2 and Table S3. Other subtests with high level of missingness are 3D Copying and Cube Copying. These three tasks required high levels of dexterity that go beyond just drawing a line on paper. Participants need to write words, pick up, and rotate small blocks. In OxVPS, the highest level of missingness were also found for the tasks requiring some dexterity or speech: cancellation, reading, and figure copy. The missingness for the Figure Copy: strategy and the Reading task: speed were likely due to an administrative error of not recording the time or strategy. In addition, it is mostly tasks near the end of RPAB and OxVPS that show higher levels of missingness, which suggests that fatigue and early termination of a session (e.g., therapy, mealtimes, or arrival of visitors) could also contribute to missingness. However, we speculate that the main reasons for missing data are due to motor impairment in the patient or aphasia.

**Table S2.**
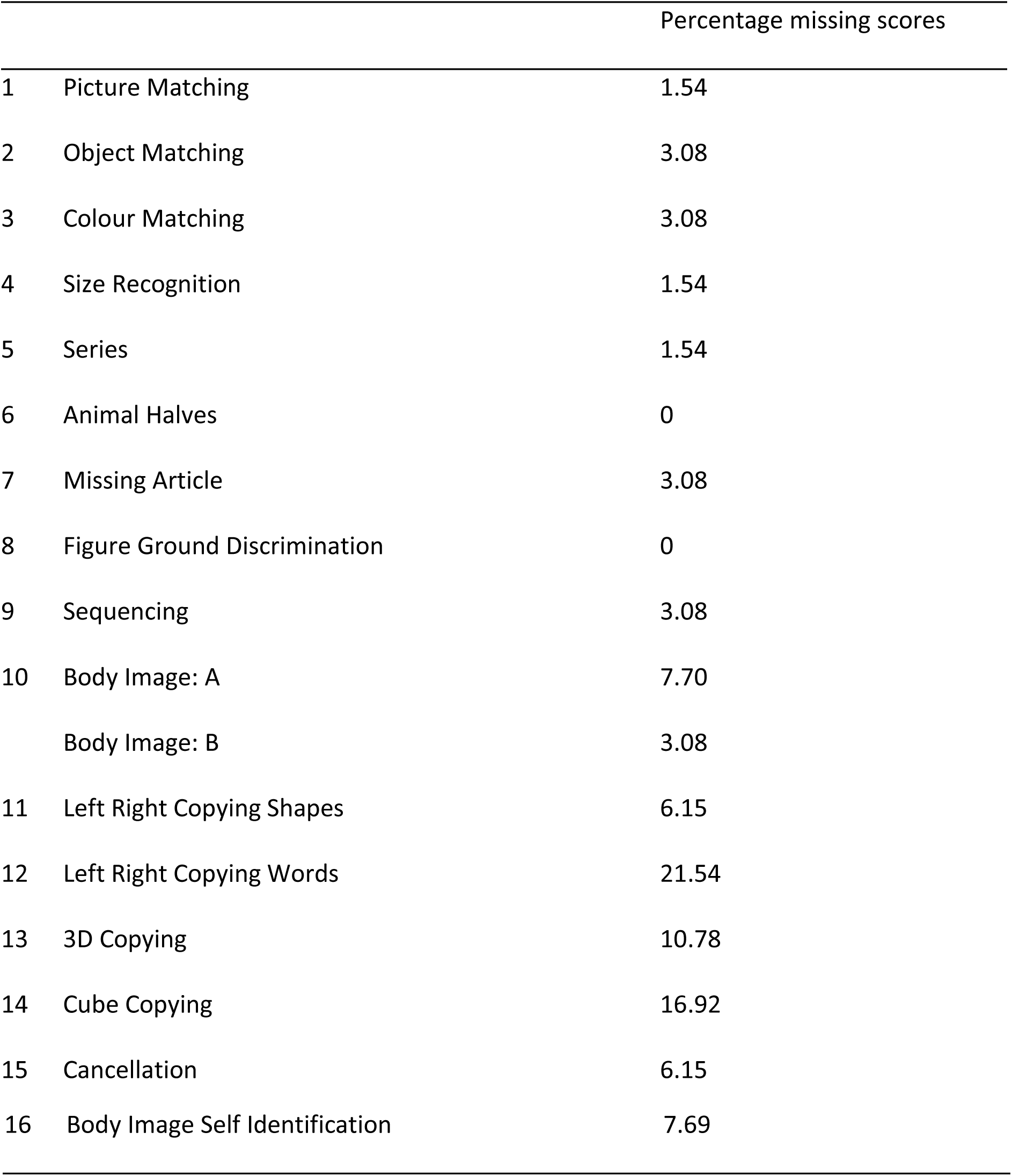
Percentage missing scores per subtest of RPAB.

**Table S3.** Percentage missing scores per subtest of OxVPS.

|  |  | Percentage missing scores |
| --- | --- | --- |
| 1 | Self-evaluation | 0 |
| 2 | Picture Naming | 0 |
| 3 | Semantic Info | 0 |
| 4 | Global Shape perception | 0 |
| 5 | Item Counting | 0 |
| 6 | Simple Feature perception | 0 |
| 7 | Face Recognition | 1.54 |
| 8 | Reading: accuracy | 4.62 |
|  | Reading: speed (word per minute) | 7.69 |
| 9 | Cancellation: space-based asymmetry | 6.15 |
|  | Cancellation: object-based asymmetry | 6.15 |
| 10 | Figure Copy: score | 3.08 |
|  | Figure Copy: strategy | 4.62 |

Missing data in demographics were not further explored, because data were only used to describe the sample, not to evaluate the effects on visual perception outcome (OxVPS and RPAB scores). Missing data in demographics is mentioned in Table 1 as ‘information not available’ and briefly discussed in the Discussion (e.g. high missingness in data on qualifications).

### Sensitivity analyses for modified Youden index

The modified Youden index was calculated for each possible cut-off point of OxVPS as Ymc=wsens∗Sens+wspec∗SpecYmc=wsens∗Sens+wspec∗Spec

With *Y_mc_* being the modified Youden index at cut-off point *c, w_sens_* and *w_spec_* as the weights for sensitivity and specificity, *Sens* as sensitivity, and *Spec* as specificity. Next the maximum *Y_mc_* is selected and at cut-off *c*, the sensitivity, specificity, positive predictive value and negative predictive values are calculated.

**Table S4.** Results of sensitivity analyses for different weight of sensitivity and specificity.

| Weight for sensitivity<br>$w_{sens}$ | Weight for<br>specificity<br>$w_{spec}$ | Cut-off<br>$c$ | Sensitivity | Specificity | PPV | NPV |
| --- | --- | --- | --- | --- | --- | --- |
| 0.8 | 0.2 | 2 | 0.87 | 0.80 | 0.93 | 0.67 |
| 0.7 | 0.3 | 2 | 0.87 | 0.80 | 0.93 | 0.67 |
| 0.6 | 0.4 | 2 | 0.87 | 0.80 | 0.93 | 0.67 |
| 0.5 | 0.5 | 2 | 0.87 | 0.80 | 0.93 | 0.67 |
| 0.4 | 0.6 | 3 | 0.68 | 0.93 | 0.97 | 0.48 |
| 0.3 | 0.7 | 3 | 0.68 | 0.93 | 0.97 | 0.48 |
| 0.2 | 0.8 | 3 | 0.68 | 0.93 | 0.97 | 0.48 |

Figures for completed cases versus the full sample

**Figure S1:**
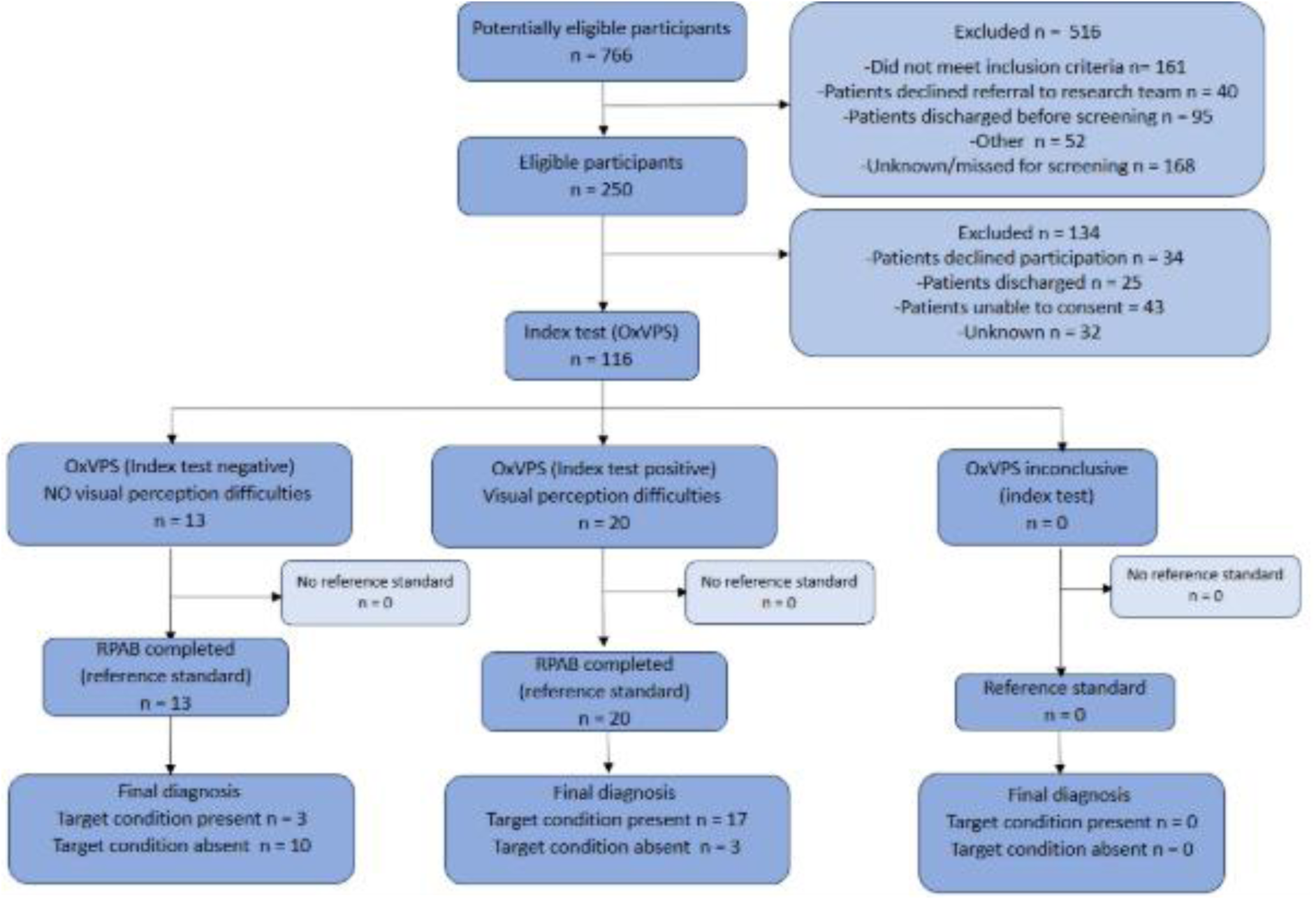
STARD flowchart for index and reference test outcomes for completed cases.

**Figure S2:**
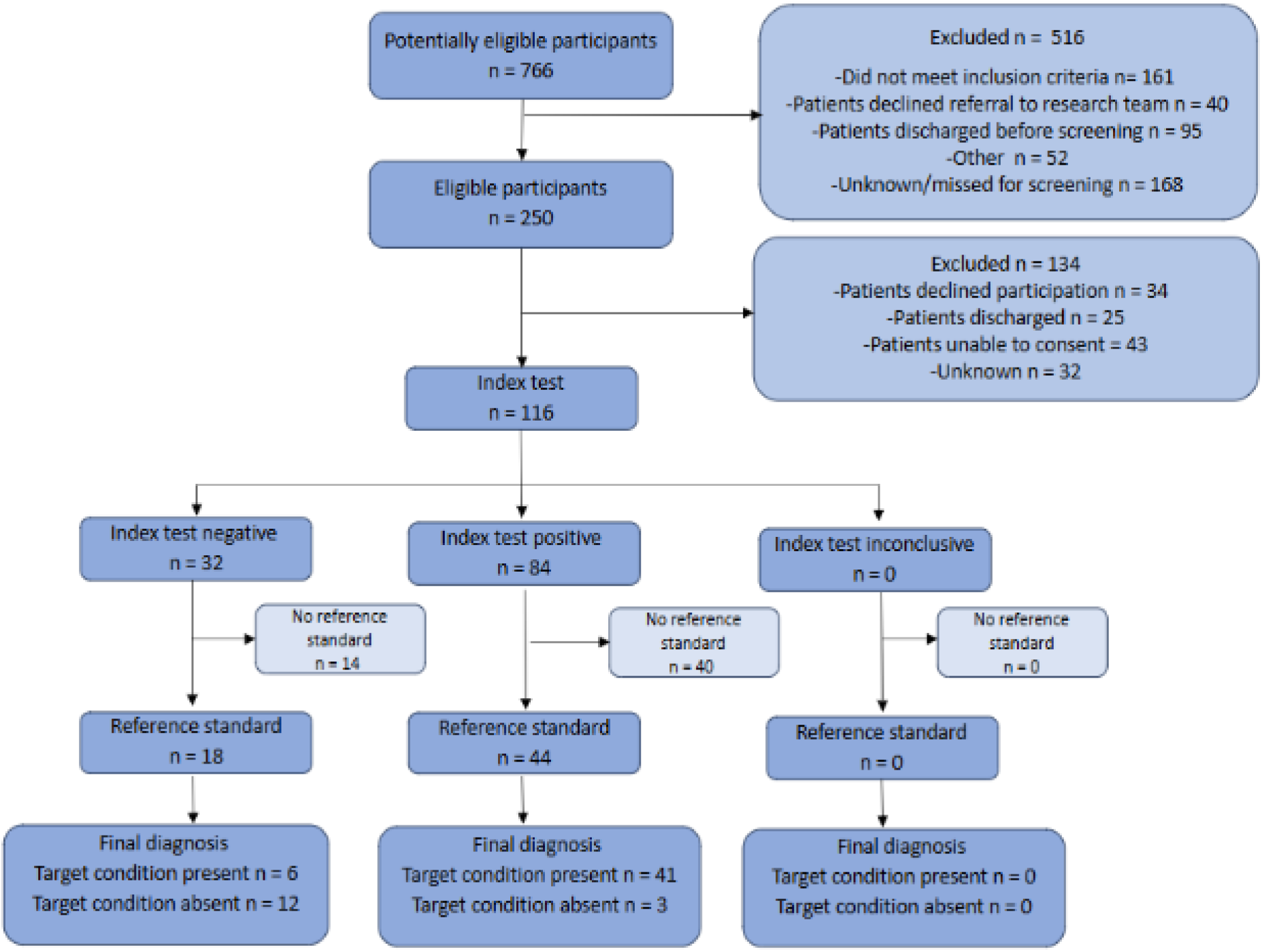
STARD flowchart for index and reference test outcomes for full sample.

